# Non-COVID-19 mortality in France, April 2020 - June 2022: reduction compared to pre-pandemic mortality patterns, relative increase during the Omicron period, and the importance of detecting SARS-CoV-2 infections

**DOI:** 10.1101/2022.11.28.22282832

**Authors:** Edward Goldstein

## Abstract

**Aims:** We compared the number of non-COVID-19 deaths between April 2020 and June 2022 to the expected number of deaths based on the patterns observed in the five years prior to the pandemic in France with the aims of (a) estimating the reduction in non-COVID-19 mortality, particularly due to reduction in the circulation of other respiratory viruses during the pandemic; (b) examining the degree to which SARS-CoV-2 infection was detected and characterized as a cause of death during different periods of the pandemic.

**Methods:** Using a previously developed regression model, we expressed weekly mortality rates in the 5-year period prior to the pandemic as a combination of influenza-associated mortality rates and baseline and a linear trend for the rates of non-influenza mortality. Estimates for the baseline and trend for non-influenza mortality together with estimates of influenza-related mortality prior to the pandemic were used to estimate expected mortality during the pandemic period.

**Results:** The number of recorded non-COVID-19 deaths between week 15, 2020 and week 26, 2022 in France was less than the expected number of deaths by 49,623 (95% CI (20364,78837)). Additionally, rates of non-COVID-19 mortality increased during the later part of the study period, with the difference between the number of non-COVID-19 deaths and the expected number of deaths during the last 52 weeks of the study period being greater than the corresponding difference for the first 52 weeks of the study period by 28,954 (24979,32918) deaths.

**Conclusions:** Our results suggest (a) the effectiveness of mitigation measures during the pandemic for reducing the rates of non-COVID-19 mortality, particularly mortality related to circulation of other respiratory viruses, including influenza (that was responsible for an annual average of 15,334 (12593,18077) deaths between 2015-2019 in France); (b) detection of a high proportion of SARS-CoV-2 infections leading to deaths in France, and characterization of those infections as the underlying cause of death. Additionally, while the increase in non-COVID-19 mortality during the later part of the study period is partly related to the temporal increase in the circulation of other respiratory viruses, there was an increase, particularly during the period of the circulation of the Omicron variant, in the proportion of hospitalizations with a SARS-CoV-2 infection in France that were coded as hospitalizations with COVID-19 (rather than COVID-19 hospitalizations), suggesting an increasing proportion of SARS-COV-2-associated deaths not being coded as COVID-19 deaths. All of this suggests the importance of timely detection of infections with SARS-CoV-2, particularly the Omicron variant (for which manifestations of disease complications are different compared to the earlier variants), and of providing the necessary treatment to patients to avoid progression to fatal outcomes.

## Introduction

Mitigation measures introduced during the COVID-19 pandemic and cross-immunity stemming from the circulation of SARS-CoV-2 viruses resulted in a significant disruption in the patterns of circulation of other respiratory viruses [1,2], with the rates of circulation for other respiratory viruses being particularly low during the winter of 2020-2021 and rebounding somewhat during the winter of 2021-2022 [1]. The reduction in the circulation of other respiratory viruses resulted in the reduction for the rates of associated mortality. Examination of mortality in England and Wales between March 2020 and June 2022 suggested that for deaths for which COVID-19 wasn’t coded as the underlying cause of death, there was a reduction of 36,7000 in the number of deaths for respiratory causes compared to averages during the previous 5 years [3]. For all deaths where the underlying cause of death was not COVID-19, excess deaths were 7,360 below the previous five-year average [3], though these estimates didn’t account for upward trends in overall mortality due to population aging and other factors. While for England and Wales, excess mortality during the pandemic was lower than the number of COVID-19 deaths, for many countries, including the US, excess mortality during the pandemic was greater than the number of COVID-19 deaths [4,5], suggesting under-detection of SARS-CoV-2 infection that resulted in various complications (not only for respiratory causes, but also for metabolic disease, circulatory causes, and other causes [3]) leading to deaths.

During the pandemic, France experienced lower cases fatality ratios (defined as the ratio between the number of deaths coded as COVID-19 deaths and the number of detected COVID-19 cases) compared to a number of other countries [6], suggesting good practices for testing and detecting cases of SARS-CoV-2 infection. While patterns of COVID-19 mortality (mortality with COVID-19 listed as the underlying cause of death) in France during the pandemic are well-characterized (with 158,539 COVID-19 deaths recorded in France by Nov. 23, 2022 [7]), patterns of non-COVID-19 mortality in France during the pandemic compared to mortality patterns in France during previous years need a better characterization. Moreover, patterns of non-COVID-19 mortality changed during the course of the pandemic in France with the increasing circulation of other respiratory viruses, and the emergence of the Omicron variant. In particular, the proportion of hospitalizations with a SARS-CoV-2 infection in France that were coded as hospitalizations with COVD-19 (rather than COVID-19 hospitalizations) increased during the period of Omicron circulation [7], suggesting an increasing proportion of SARS-COV-2-associated deaths not being coded as COVID-19 deaths. The relative risk for complications, including hospital admission and death in adults is lower for the Omicron variant compared to the Delta variant, though those relative risks vary with age, with the relative risk for severe outcomes, including death for Omicron vs. Delta being greatest for the oldest adults [8,9]. This is related to differences in disease manifestation for Omicron infections vs. Delta infections for both ED admissions [10] and hospitalizations [11], which is also related to the above changes in coding for hospitalized patients with a detected Omicron infection in France [7]. Better understanding is needed regarding how differences in disease manifestation for complications stemming from Omicron infections compared to earlier SARS-CoV-2 variants are related to differences in treatment of Omicron infections, and to patterns in non-COVID-19 mortality (some of which is Omicron-associated, though not recognized as COVID-19 mortality).

In this study, we examined how rates of non-COVID-19 mortality during different stages of the pandemic in France compared to patterns in mortality during the 2015-2019 period. For the pre-pandemic period, we applied a previously developed regression model [12] to express mortality rates between 2015-2019 as a combination of influenza-associated mortality rates and baseline and a linear trend for the rates of non-influenza mortality. The estimates for the baseline and trend for non-influenza mortality prior to the pandemic together with the estimates of influenza-related mortality prior to the pandemic led to the estimation of expected mortality during the pandemic period. That expected mortality was then compared to rates of non-COVID-19 mortality in France during different periods between April 2020 and June 2022 (with the end period for the study chosen as June 2022 due to high levels of mortality due to a heat wave in July 2022 in France [13]).

## Methods

### Data

Data on the daily number of deaths for all causes in France starting 2015 are available from [14], whereas data on the daily number of COVID-19 deaths are available from [6]. Data on population in France are available from [15]. Data on weekly numbers of influenza-like illness (ILI) consultations in metropolitan France are available from the French sentinel surveillance [16]. Sentinel data on testing of respiratory specimens for the different influenza subtypes are available from WHO FluNET [17].

### Statistical model for the 2015-2019 period

We use a previously developed model that combined data on syndromic surveillance with data on virologic surveillance to estimate rates of mortality associated with the major influenza subtypes in the US [11]. Subsequently, that method was applied to the estimation of influenza-associated mortality in other countries [18-20], including the EU population [20].

Not all ILI consultations in the sentinel data [16] correspond to influenza infections, and those that do, correspond to infection with different influenza subtypes. For each influenza subtype (e.g. A/H3N2), we define an indicator for the incidence of that subtype on week *t* (e.g. *A/H3N2*(*t*)) as

*A/H3N2*(*t*) = Rate of ILI consultations per 100,000 persons on week *t* in [16] * Percent of respiratory specimens on week *t* in [17] positive for influenza A/H3N2 (1)

To relate the incidence indicators for the major influenza subtypes to weekly rates of all-cause mortality per 100,000 persons in France, we note that age distribution of influenza cases changes with the appearance of antigenically novel influenza strains, and this changes the relation between rates of influenza-associated ILI (incidence indicators in eq. 1) and rates of influenza-associated mortality. The relevant antigenic changes for our study period were (a) the appearance of an antigenically/genetically novel A/H3N2 strain during the 2014-2015 influenza season [21]; the appearance of the novel influenza B/Yamagata strain during the 2017-2018 season [20]; the 2018-2019 A/H3N2 epidemic strains belonging to several clades and exhibiting different immunity profiles for different birth cohorts [22]. Correspondingly, for the model relating A/H3N2 circulation to associated mortality, we split the A/H3N2 incidence indicator into three: *A/H3N2*_1_, equaling the A/H3N2 incidence indicator for the period from Jan. 2015 to Sep. 2015, and equaling to 0 for later weeks; *A/H3N2*_2_, equaling the A/H3N2 incidence indicator for the period from Oct. 2015 to Sep. 2018, and equaling to 0 for other weeks, and *A/H3N2*_3_, equaling the A/H3N2 incidence indicator for the period from Oct. 2018 to Jan. 2020, and equaling to 0 for other weeks. Similarly, we split the B/Yamagata incidence indicator into two, corresponding to the periods before and starting the 2017-2018 B/Yamagata epidemic. Finally, we note that it takes 1-2 weeks between influenza illness and influenza-associated mortality [11].

Correspondingly, we relate the mortality rate *M*(*t*) on week *t* to the *shifted* incidence indicators, e.g.

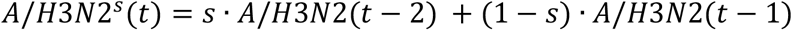

where the parameter *s* (common for all incidence indicators) is chosen to minimize the R-squared for the model fit. The regression model that we use is:

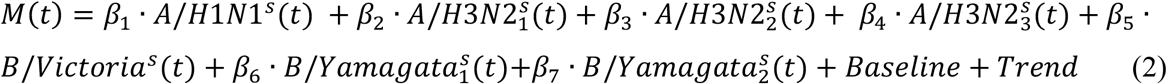

Here, Baseline represents weekly rates of mortality not associated with influenza circulation, and *Baseline*(*t*) is modelled to have annual periodicity in week *t*. We use periodic cubic splines to model the baseline mortality rates whose shape is unknown [11]. The trend *Trend*(*t*) is modelled as a linear polynomial in week *t* (see Discussion). Finally, to account for the autocorrelation in the noise, we use a bootstrap procedure (resampling the noise on different weeks) to estimate the confidence bounds for various quantities evaluated in the model [11].

### Statistical analysis for the pandemic period

Baseline and the linear trend for mortality rates not associated with influenza are extended into the pandemic period (Figure 1) to model expected rates of non-influenza mortality during the pandemic period. Average annual estimates for the number of influenza-associated deaths for the 2015-2019 period are used to model the expected rates of influenza-associated mortality during each season of the pandemic period. Bootstrap sample for all the parameters in eq. 2 is used to estimate means and 95% credible intervals for the differences between observed rates of non-COVID-19 deaths and rates of expected mortality during different periods between week 15, 2020 (beginning on April 6, 2020) and week 26, 2022 (beginning on June 27, 2022).

**Figure 1:**
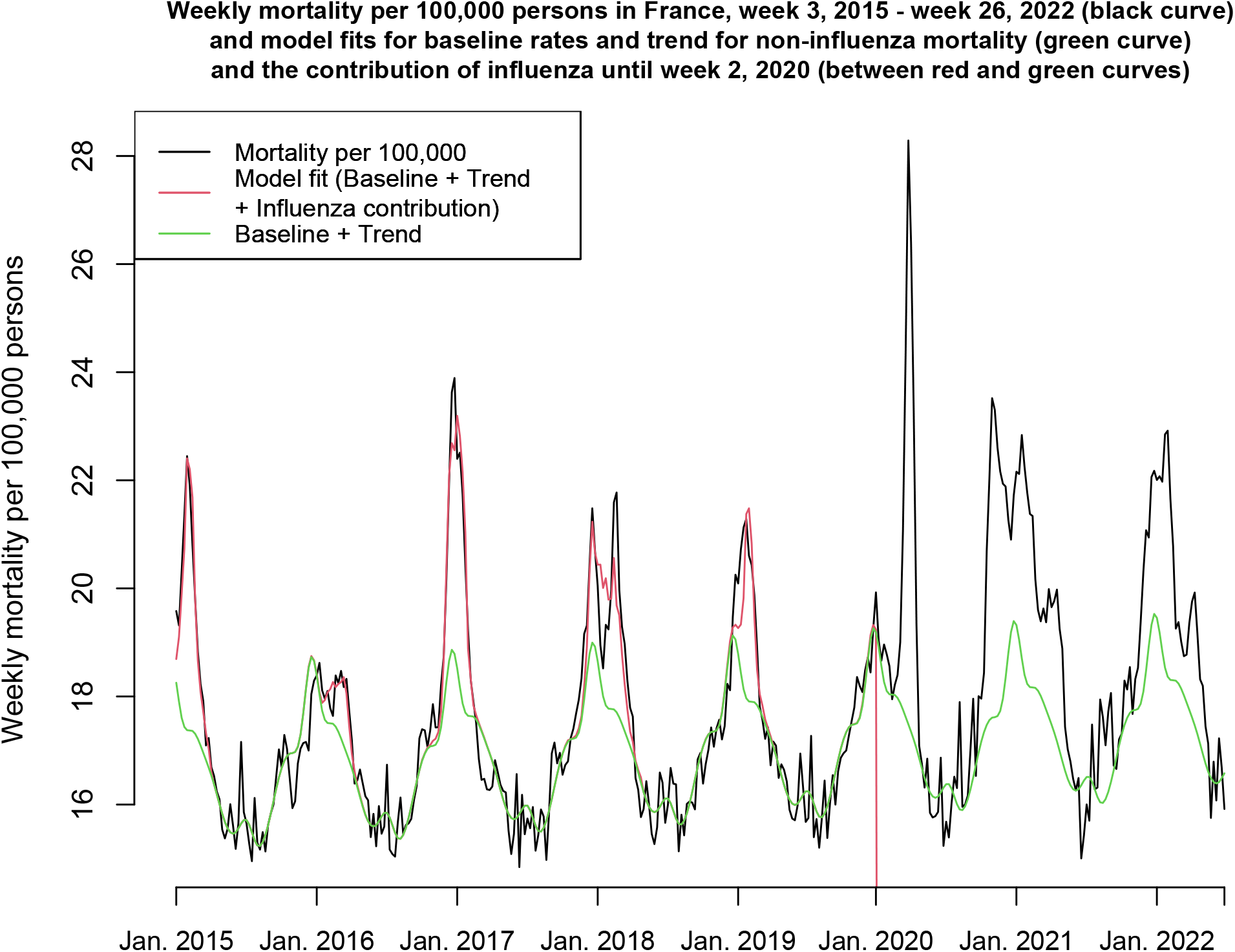
Weekly mortality rates per 100,000 persons in France between week 3, 2015 – week 26, 2022, and model fits for baseline rates and trend (green curve) and the contribution of influenza (between red and green curves) for the period between week 3, 2015 and week 2, 2020.

## Results

Figure 1 plots the weekly mortality rates per 100,000 persons in France between week 3, 2015 – week 26, 2022, as well as the results of the fit for the model given by eq. 2. for the period between week 3, 2015 and week 2, 2020. The model fit is generally temporally consistent except for the 2017-2018 season during which the circulating A/H1N1 and B/Yamagata strains had a different age distribution (Discussion). Baseline and the linear trend for the mortality rates not associated with influenza are extended into the pandemic period (through week 26, 2022, Figure 1).

Figure 2 plots the weekly rates of non-COVID-19 mortality between week 15, 2020 and week 26, 2022 in France, as well as the estimates for the baseline and trend for the rates of non-influenza mortality. Figure 2 shows an increase in non-COVID-19 mortality during the later part of the study period. The difference between the number of non-COVID-19 deaths and the expected number of deaths during the last 52 weeks of the study period was greater than the corresponding difference for the first 52 weeks of the study period by 28,954 (24979,32918) deaths.

**Figure 2:**
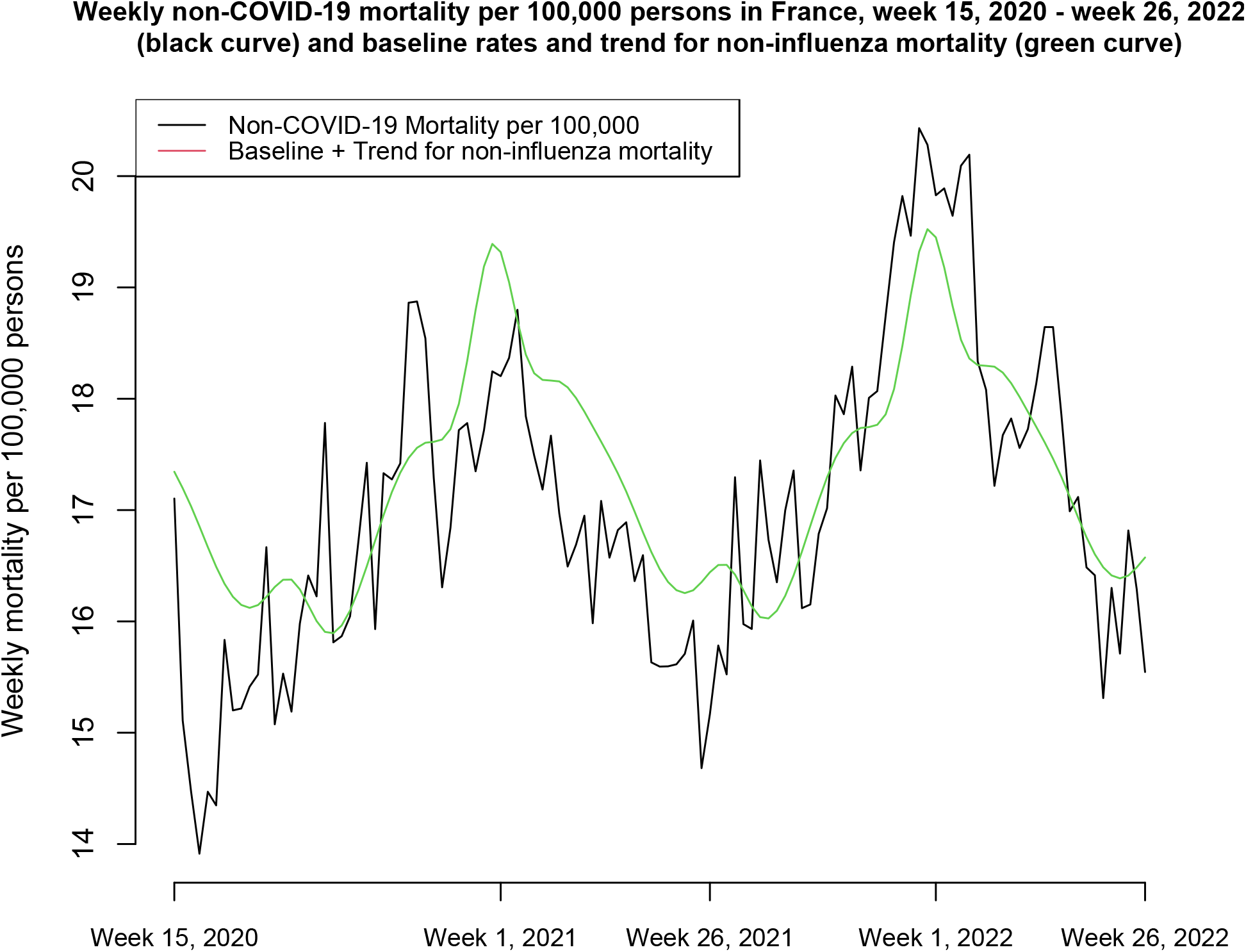
Weekly rates of non-COVID-19 mortality between week 15, 2020 and week 26, 2022, and the estimates for the baseline and trend for the rates of non-influenza mortality.

Table 1 provides estimates for the various quantities relevant to the analyses in the paper. Between week 3, 2015 – week 2, 2020 there was an annual average of 15,334 (12593,18077) influenza-associated deaths. The expected number of non-influenza deaths (given by the combination of the baseline and trend for non-influenza mortality) between week 15, 2020 – week 26, 2022 was 1,366,959 (1337438,1396528). The total expected number of deaths (non-influenza deaths + expected influenza-associated deaths) between week 15, 2020 – week 26, 2022 was 1,397,627 (1368368,1426841). The number of recorded non-COVID-19 deaths between week 15, 2020 – week 26, 2022 was 1,348,004. The difference between the expected number of deaths and the number of recorded non-COVID-19 deaths between week 15, 2020 – week 26, 2022 was 49,623 (20364,78837).

**Table 1:** Estimates for the average annual number of influenza-associated death between 2015-2019, the expected number of deaths between week 15, 2020 – week 26, 2022, and the difference between the expected number of deaths and the number of recorded non-COVID-19 deaths between week 15, 2020 – week 26, 2022.

|  |  |
| --- | --- |
| Average annual number of influenza-associated deaths between 2015-2019 | 15,334 (12593,18077) |
| Expected number of non-influenza deaths between week 15, 2020 – week 26, 2022 | 1,366,959 (1337438,1396528) |
| Total expected number of deaths between week 15, 2020 – week 26, 2022 | 1,397,627 (1368368,1426841) |
| Number of recorded non-COVID-19 deaths between week 15, 2020 – week 26, 2022 | 1,348,004 |
| Difference between the expected number of deaths and the number of recorded non-COVID-19 deaths, week 15, 2020 – week 26, 2022 | 49,623 (20364,78837) |

## Discussion

Mitigation measures introduced during the COVID-19 pandemic and cross-immunity stemming from the circulation of SARS-CoV-2 viruses resulted in a significant disruption in the patterns of circulation of other respiratory viruses [1,2], and reduction in the rates of respiratory non-COVID-19 mortality, as well as mortality for certain other causes compared to pre-pandemic patterns [3]. The effect of the pandemic on excess mortality for all causes and its relation to the number of COVID-19 related deaths varied between the different countries [3-5], with some countries, including the US, reporting significantly higher excess mortality rates compared to rates of COVID-19 deaths. The latter is related to under-detection of SARS-CoV-2 infections in associated complications leading to fatal outcomes for different causes (including not only respiratory causes, but also metabolic disease, circulatory causes, and other causes [3]). During the pandemic, France experienced lower case fatality ratios (the ratio between the number of COVID-19 deaths and the number of detected COVID-19 cases) compared to a number of other countries [6], suggesting good practices for testing and detecting cases of SARS-CoV-2 infection. This suggests that evaluating patterns of non-COVID-19 mortality during the pandemic period in France in relation to patterns in all-cause mortality prior to the pandemic should be useful for estimating the reduction in non-COVID-19 mortality compared to pre-pandemic mortality, and for examining the degree to which SARS-CoV-2 infection was detected and characterized as a cause of death during different periods of the pandemic in France. We used a previously developed model [11] to estimate the baseline and trend for the rates of non-influenza mortality between 2015-2019, as well as the rates of influenza-associated mortality between 2015-2019 -- see more on influenza-associated mortality in France, including its relation to vaccination and antiviral use in [23]. Those estimates were then used to estimate expected mortality during the pandemic period and compare it to the recorded non-COVID-19 mortality. We found that the number of recorded non-COVID-19 deaths between week 15, 2020 and week 26, 2022 in France was less than the expected number of deaths by 49,623 (20364,78837). Reduction in non-COVID-19 mortality, particularly though the reduction in the circulation of other respiratory viruses, including influenza (that was responsible for an annual average of 15,334 (12593,18077) deaths between 2015-2019 in France) had somewhat mitigated the overall mortality burden during the pandemic (with 158,539 COVID-19 deaths recorded in France by Nov. 23, 2022 [7]). However, reduction in non-COVID-19 mortality was not uniform during different periods of the pandemic, with the difference between the expected number of deaths and the number of non-COVID-19 deaths declining during the later part of the study period (July 2021-June 2022) compared to the earlier part of the study period. That increase in the number of non-COVID-19 deaths is related to both the increase in the circulation of other respiratory viruses, including influenza, but also to the emergence of the Omicron variant. In particular, the proportion of hospitalizations with a SARS-CoV-2 infection in France that were coded as hospitalizations with COVD-19 (rather than COVID-19 hospitalizations) increased during the period of Omicron circulation [7], which is related to differences in disease manifestation for Omicron infections vs. Delta infections in both ED admissions [10] and hospitalizations [11]. All of this suggests that an increasing proportion of SARS-COV-2-associated deaths are not being coded as COVID-19 deaths during the Omicron period. It is therefore important to detect infections with SARS-CoV-2, particularly the Omicron variant, and to provide the necessary treatment to patients (particularly the oldest individuals, for whom the severity of the Omicron variant, not just in absolute terms, but also relative to the earlier SARS-CoV-2 variants is greatest among all age groups of adults [8,9]) avoid progression to fatal outcomes.

Our results have some limitations. Influenza surveillance data in France pertains to mainland France [16], whereas we’ve used data on mortality for the whole of France. Additionally, sentinel data on testing for viral specimens [17] has a moderate sample size and may not represent all of France. We note that influenza epidemics exhibit a great deal of temporal synchrony [24,25] which should help address the above limitations. Despite the fact that we split some of the influenza subtype incidence indicators into several time periods, where might still be temporal variability in the relation between the incidence indicators used in this paper and rates of associated mortality. For example, while model fits are generally temporally consistent (Figure 1), the model fit for the mortality data for the 2017-2018 season is worse compared to other influenza seasons, which might be related to the fact that the influenza subtypes that circulated during that season (A/H1N1 and B/Yamagata) have different age distributions feeding into one ILI data stream. Finally, we use a linear trend for the rates of non-influenza mortality between 2015-2019. Using a quadratic polynomial for the trend ([23]) and extending it to the pandemic period results in higher estimates for the expected rates of non-influenza mortality during the pandemic period, and a bigger difference between the expected number of deaths and the number of recorded non-COVID-19 deaths between week 15, 2020 and week 26, 2022 in France. While there is some uncertainty regarding how baseline and trend for the rates of non-influenza mortality would have extended until June 2022 in the absence of the pandemic, our use of the linear model for the trend likely provides conservative estimates for the difference between the expected mortality rates and the recorded non-COVID-19 deaths between week 15, 2020 and week 26, 2022 in France.

## Conclusions

Our results suggest the effectiveness of mitigation measures during the pandemic for reducing the rates of non-COVID-19 mortality, particularly mortality related to the circulation of other respiratory viruses, including influenza, with the reduction in non-COVID-19 mortality rates somewhat mitigating the overall mortality burden during the pandemic in France. Our results also suggest the detection of a high proportion of SARS-CoV-2 infections leading to deaths in France and characterization of those infections as the underlying cause of death. The latter has changed somewhat during the Omicron period [7], which is related to the increase in non-COVID-19 mortality in France during the Omicron period, as well as to changes in disease manifestation and in coding for complications stemming from Omicron infections compared to the earlier SARS-CoV-2 variants [7,10,11]. Those changes are not uniform across the different age groups -- for the oldest individuals, the severity of the Omicron variant relative to the earlier SARS-CoV-2 variants is greatest among all age groups of adults [8,9]. This suggests the importance of timely detection of infections with SARS-CoV-2, particularly the Omicron variant, and of providing the necessary treatment to patients (especially the oldest individuals) to avoid progression to fatal outcomes.

## Data Availability

This study is based on de-identified, aggregate data available through refs. 6,14,15,16,17

https://covid19.who.int/

https://www.insee.fr/fr/statistiques/4931039?sommaire=4487854#tableau-figure1

https://www.insee.fr/fr/statistiques?debut=0&theme=1

https://www.sentiweb.fr/france/fr/?page=table

https://app.powerbi.com/view?r=eyJrIjoiNjViM2Y4NjktMjJmMC00Y2NjLWFmOWQtODQ0NjZkNWM1YzNmIiwidCI6ImY2MTBjMGI3LWJkMjQtNGIzOS04MTBiLTNkYzI4MGFmYjU5MCIsImMiOjh9

